# Have we forgotten our obligation to train health workers on disability? A policy analysis in sub-Saharan Africa

**DOI:** 10.1101/2024.04.30.24306648

**Authors:** Sara Rotenberg, Veronika Reichenberger, Tracey Smythe

## Abstract

People with disabilities generally have worse health outcomes than people without disabilities, leading to a 10–20-year difference in life expectancy. Research on the barriers to accessing health care frequently points to the role of health workers’ attitudes and lack of training to provide high quality health care to people with disabilities. Current training initiatives are unsystematic and limited to specific cadres or institutions. Yet, many countries that have adopted the UN Convention on the Rights of Persons with Disabilities likely have legal obligations to train health workers on disability in these laws. The purpose of this paper was to systematically explore the laws and policies in sub-Saharan Africa to understand how countries should be training their health workers. We searched WHO MiNDBANK and UN websites for disability laws and policies. We systemically extracted information across 11 domains: 1) requirements, 2) training objectives, 3) training cost, 4) training duration, 5) competencies covered, 6) educational stage, 7) training methods, 8) impairment-specific, 9) cadres, 10) benefits for attendance, and 11) monitoring and evaluation plans. 53 documents in English, French, and Portuguese from 32 countries were eligible for inclusion, while 16 countries had no disability laws or policies. Of the documents included, 24 (45%) did not have any mention of health worker training, while 17 (32%) recommended and 10 (19%) required health worker training. Most laws had no further specifications to describe training, though more robust laws and policies had information on the budget allocation, competencies, educational stage, and cadres included. Several countries in sub-Saharan Africa do have disability laws that require health worker training, and more countries should be including health worker training in their curricula to comply with their national laws.

**Key messages (2-4):**

- Out of the 48 countries included, 16 had no disability laws, policies, or strategies in eligible databases. While most countries adopted disability policies following the implementation of the UN CRPD in 2006, there remains a notable absence of current and comprehensive disability legislation in many areas, affecting the scope and effectiveness of disability training for health workers.
- Nearly half of the documents reviewed across 48 countries did not mention disability training for health workers within their national disability laws or policies. Where training was mentioned, it varied significantly, with some countries recommending or mandating training, but often limiting it to specific health worker groups, which might not comprehensively cover all healthcare providers who encounter disabled patients.
- Malawi, Lesotho, and Rwanda are notable for their detailed training objectives and evaluation plans within their disability policies. These countries provide examples of more proactive approaches, focusing on specific training needs such as sign language and the inclusion of budgeting for training implementation.
- Despite the presence of laws and policies, there is often a lack of detailed implementation and monitoring plans, which limits the effectiveness of these policies. The study highlights the need for laws and policies to be accompanied by specific, actionable, and funded plans to ensure that disability training for health workers is not only mandated but also effectively implemented.

**Reflexivity statement:** The authors of this paper include 3 women who are experts on disability and health, two of whom are from low- and middle-income countries. One author is disabled and another is from sub-Saharan Africa.

## Background

Globally, there are 1.3 billion people with disabilities, many of whom experience barriers in accessing health care (World Health Organization, 2022b), leading to a 10-20 year gap in life expectancy amongst this population. (The Missing Billion Initiative and Clinton Health Access Initative, 2022) Health worker attitudes, stigma, and poor training is frequently cited as one of the key barriers for people with disabilities to access healthcare. (Rotenberg et al., 2022, Hashemi et al., 2020) For example, a recent study in the US showed that the majority of doctors feel unconfident in providing care to people with disabilities (Iezzoni et al., 2021) and most want more training on disability.(Marzolf et al., 2022)

Over the past two decades, international organisations and laws have also increasingly called for more disability training for health workers through global pledges and commitments. For example, the World Health Organization includes several specific recommendations on training health workers as part of their 40 actions on achieving health equity for disabled people, released in December 2022.(World Health Organization, 2022a) Furthermore, Article 25(d) of the UN CRPD specifies health worker training as a means of improving health equity and quality of care for disabled people.(United Nations, 2006) Regionally, the African Union has protected the rights of disabled people through the African Disability Protocol, which mandates several stipulations in regards to the right to health. Several clauses of Article 17 of the African Disability Protocol address stigma, discrimination, and health worker training, informing the importance of this topic in African countries’ disability laws.(African Union, 2018) Accordingly, several countries in sub-Saharan Africa include disability training in their national disability laws, committing to this central component of improving health for disabled people. Countries, therefore, have a domestic and international legal obligation to deliver disability training for health workers by ratifying these treaties and protocols. However, there remains a persistent gap between ratification and implementation.

Progress on disability-inclusion is important to contextualize within legal institutions, because disability-inclusion may only begin after laws and policies have changed to reflect increasing acceptance of disabled people.(Goodley, 2017) That is, underlying negative attitudes and actions towards disabled people have often changed after advocacy and activism have focused on enactment of anti-discrimination laws and policies as one of the main mechanisms for improving disability-inclusion.(Goodley, 2017, Shakespeare, 2014) Solidifying these efforts with laws becomes a particularly important tool for enacting and enforcing these changes, since there is often a void or resistance to enact elements of anti-discrimination laws. This socio-legal approach to social change demonstrates the importance of understanding the legal and policy context for disability inclusion in health worker training. Legislation on training may be an opportunity to integrate disability into curricula or competency requirements. By enacting a law that mandates training for healthcare workers on disability, it ensures that health professionals are equipped with the necessary knowledge and skills to understand and address the needs and challenges faced by disabled patients. This training may encompass disability awareness, communication strategies, providing culturally competent care, and ensuring accessible health facilities.(Shakespeare and Kleine, 2013, Azizatunnisa et al., 2023) Ultimately, such a law aims to promote inclusivity and equitable health services for disabled people, fostering a more compassionate and accommodating healthcare environment for all patients.

This makes it important to examine how disability law deals with disability training for health workers, if at all. Similarly, examining how national disability strategies have incorporated disability training can be insightful for how countries implement policy in practice and how individual governments have conceptualised their obligations under UN CRPD. To date, most analysis of disability laws has focused on how these laws support inclusive education for disabled children.(Bose and Heymann, 2020) However, very little research has examined the right to health within these laws or determining countries’ obligations to deliver health worker training.

## Methods

Using the READ approach (Dalglish et al., 2020) for document analysis, which involves systemically selecting and extracting data according to a framework, we examined the laws and policies of 48 countries in sub-Saharan Africa. The laws, policies, and strategies were sourced from several pre-determined reference frames that were determined to have comprehensive records of existing disability laws: WHO MiNDBANK (More Inclusiveness Needed in Disability and Development) and UN Department of Economic and Social Affairs (UN DESA) webpages. Additionally, authors familiar with in-country contexts were able to provide documents. First, we searched for each country’s page on the WHO MiNDBank (World Health Organization, 2023), which includes countries’ national policies, strategies, and laws on mental health, substance abuse, disability, general health, human rights, and development. This database was then cross-referenced with the UN DESA Disability Strategies and Action Plans by Country webpage (United Nations Department of Economic and Social Affairs, n.d.-b), as well as their Disability Laws and Acts by Country webpage (United Nations Department of Economic and Social Affairs, n.d.-a). These existing databases provided a standardized way of accessing recorded laws, policies, and strategies, to reduce bias from using other search engines unsystematically or encountering potential language or geographic biases. There were no date or language restrictions applied. The databases did not provide date of updates, but since the databases were cross-referenced, they were assumed to be up to date.

Each document was saved on the date of access and reviewed by two authors, including documents in French and Portuguese. An extraction framework was developed to count mentions of 11 specific aspects of training, including: 1) legal requirement 2) costs, 3) time, 4) competencies, 5) staging (i.e., when training is delivered in curricula), 6) training methods, 7) impairment type, 8) cadres, 9) benefits of completion, 10) objectives, and 11) evaluation and monitoring. These categories were developed using the key elements of the trainings from a systematic review of interventions to train health workers about disability.(Rotenberg et al., 2022) Data were then extracted for each aspect of the framework. An Excel-based extraction tool was used to summarise information. Finally, the extracted data were analysed by documents and by country (after selecting the most recent or highest-scoring document) to infer trends in inclusion of disability training for health workers in disability laws, policies, and strategies.

## Results

Of the 48 countries included in the sample, 16 had no disability laws, policies, or strategies in any of the databases.(World Health Organization, 2023, United Nations Department of Economic and Social Affairs, n.d.-a, United Nations Department of Economic and Social Affairs, n.d.-b) In total, 53 documents were eligible for inclusion across 32 countries. Most countries had disability policies from post-UNCRPD implementation (i.e., 2006), while four had only documents from before 2000.(Government of the Democratic Republic of the Congo, 1973, Government of Madagascar, 1998, Government of the Republic of Botswana, 1996, Government of the Republic of Gabon, 1996) Malawi and South Africa had the most documents at four each, while most other countries had a disability law only.

In terms of content, nearly half the documents (n=24, 45%) and countries (n=14, 44%) had no mention of disability training for health workers within their national disability laws or policies. Most documents recommended disability training (n=17, 32%) or required it (n=10, 19%). By country, most had at least one document that recommended training (n=11, 34%) while several had it as mandatory (n=7, 21%). Laws and policies included a mean of 1.4 items from the framework (SD = 1.8) per document and a mean of 1.7 items per country (SD = 1.9), with most signalling the cadres that should be trained and the educational staging for training (Table 1). Almost all countries mentioned the need to train health workers in the curriculum or through continuing professional development (CPD) for those already qualified. For example, in The Gambia, the law specifically mentioned integrating disability training into health programmes’ curricula and CPD for all current health and rehabilitation staff. (Government of The Gambia, 2009) Ghana’s laws highlighted the importance of including training on general health and rehabilitative care for disabled people within training for health professionals and health care programs. (Government of Ghana, 2006) Sign language was highlighted as a competency that should be included in training in Lesotho, Malawi, and Uganda, but only Lesotho’s policy specifically included that health service providers such as nurses, doctors, and HIV testing and counselling (HTC) counsellors should be trained in sign language. (Government of the Kingdom of Lesotho, 2011) While most countries included broad cadres, such as “all health workers”, others adopted a narrower scope, limiting eligibility to specific groups such as rehabilitation staff or allied health professionals, as demonstrated by Ethiopia. (Government of the Federal Democratic Republic of Ethiopia, 2011) Policy and law details are outlined in Supplementary Table 1.

**Table 1:**
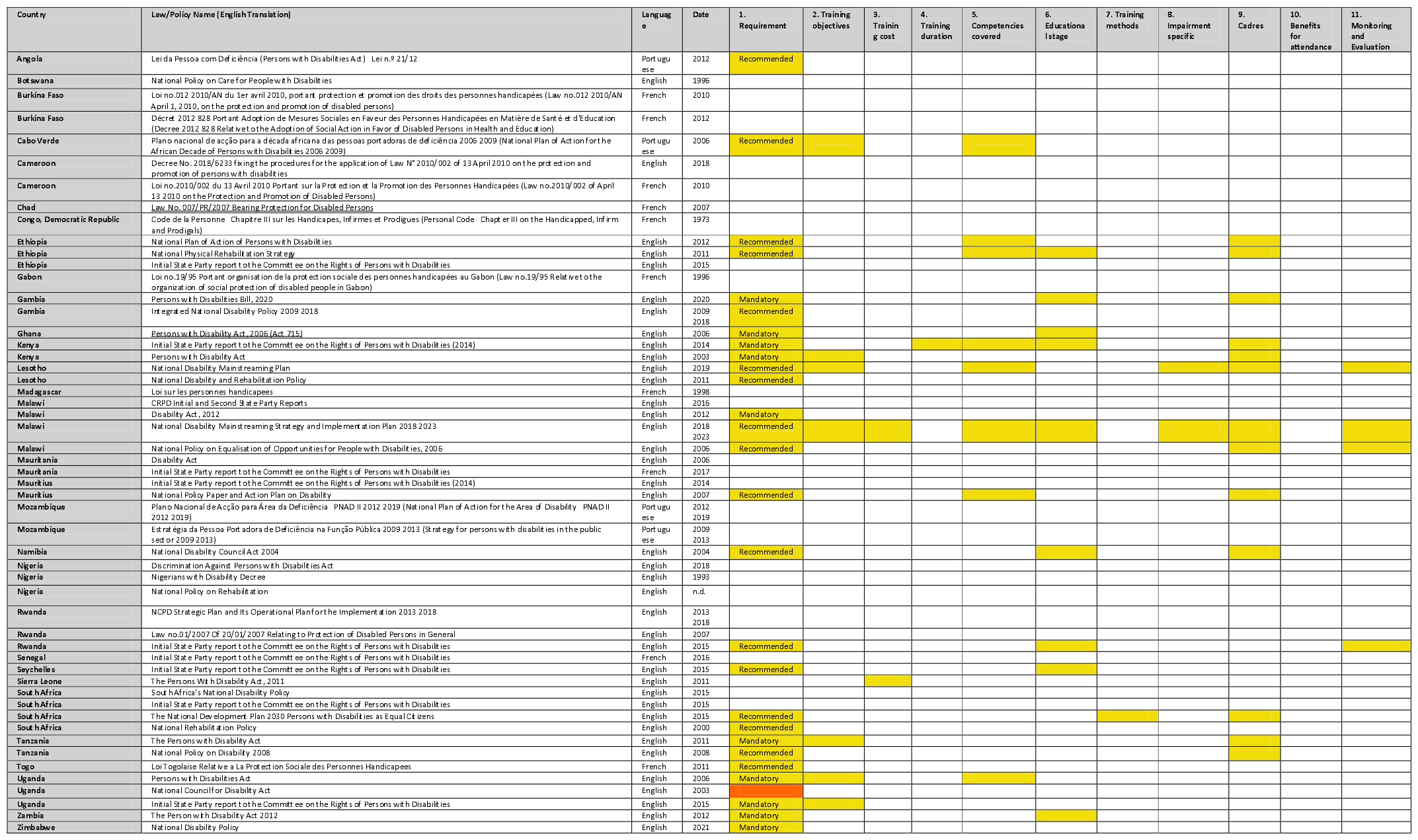
Summary of Disability Law and Policy Contents in 32 countries in sub-Saharan Africa (yellow-included)

Most laws and policies listed knowledge and skills acquisition as a primary objective of the training, but Lesotho and Uganda included sign language skills as the goal of training. Only two countries’ policies included a budget or cost of training; Malawi’s National Disability Mainstreaming Strategy and Implementation Plan 2018-2023 provided a specified budget (50 million MK; nearly £50,000 in 2018) and Sierra Leone’s Persons with Disabilities Act, 2011 stated the government would contribute to the budgets of institutions’ that trained health workers on disability. Kenya’s CRPD report was the only document to mention the length of training for rehabilitation staff (3 years), while South Africa’s The National Development Plan 2030 Persons with Disabilities as Equal Citizens policy was the only document to state methods and highlight the need for training to be led by people with disabilities themselves. Furthermore, few countries mentioned specific impairment types, except for Malawi (albinism)(Government of Malawi, 2018) and Lesotho (people with intellectual disabilities and d/Deaf and hard of hearing individuals). (Government of the Kingdom of Lesotho, 2011)

Malawi, Lesotho, and Rwanda also had plans within the policies to evaluate their laws. For example, Malawi stated the country would aim to have three new curricula for health personnel; 41 physiotherapists trained; 100 rehabilitation technicians trained (Government of Malawi, 2018) while Lesotho’s plan was broader and focused on providing sign language skills to health service providers including nurses, doctors, HTC counsellors, and other health workers in 5 years.(Ministry of Social Development Lesotho, 2019) In Rwanda, the UN CRPD report stated 600 staff had been trained in 30 districts over three days in 2013. (Government of the Republic of Rwanda, 2013) These specifics were not found in any other countries’ policies or laws, making them the clearest examples of commitment and monitoring among all the laws discussed in this section.

Malawi had the greatest number of laws, policies, UN CRPD reports, and strategies (n=8), as well as substantial details about the specifics of training. For example, Malawi’s National Disability Mainstreaming Strategy and Implementation Plan 2018-2023 had the highest score of all documents analysed, with specific details in 73% of categories included in the framework (n = 8). This document emphasized disability training for health workers since access to health services was selected as one of the priority areas for the government. Accordingly, the policy specifies the budget and specific competencies (sign language, causes of disability, early intervention, assessment, and referral services), disabilities (albinism), health cadres (physiotherapists, orthopaedics, Dermatologists and ophthalmologists, HTC service providers), and objectives of the training (highlighted above).

## Discussion

Our findings highlight both the progress and the gaps in disability inclusion within healthcare systems across sub-Saharan Africa. We examined disability laws, policies, and strategies across 48 countries, revealing significant disparities in their inclusion and content. Notably, 17 countries lacked any such legislation. Most countries implemented disability policies post-UN CRPD (2006), indicating a need for international standards and frameworks to promote and guide the development of inclusive disability policies and strategies worldwide. Additionally, the lack of up-to-date laws make it challenging to assess the state of disability rights, particularly concerning the health system’s efforts to adequately train health workers on treating disabled patients. Moreover, nearly half of the countries did not address disability training for health workers in their laws or policies. While many countries recommended or required such training, the scope varied, with some focusing on broad cadres while others specified certain groups. Few countries allocated budgets for training or mentioned evaluation plans for their laws. Malawi stood out for its comprehensive approach, including detailed training objectives, budget allocation, and plans for evaluation.

While these documents provided some evidence that there has been some attention to training to improve health equity for disabled people, the majority of these inclusions were minimally modified articles of UN CRPD. This boiler-plate approach can provide some basis for action. For example, in Ghana, the language of their Disability Act is similar to UN CRPD, yet there have been various successful individual disability trainings and interventions.(Ganle et al., 2021) However, these trainings have not been integrated into curricula systemically, even though that could be expected from implementing such a law. Therefore, having further documents that delineate plans, budget, and evaluation (i.e., Malawi’s mainstreaming strategy) can support actual disability training integration into the health worker education system.

Furthermore, most of the countries only listed information about the requirements for training, rather than in-depth content. The absence of disability action plans or policies adjacent to the laws may suggest that the laws themselves may not further these objectives. Whilst laws stated that disability training was required, this mandate does not necessarily translate to high-quality disability training. Countries with more detailed training descriptions within their laws may not necessarily produce better trained health workers. For example, Lesotho’s focus on sign language competency can be exceptionally beneficial for d/Deaf and hard of hearing patients, but this requires that health education systems have the capacity to teach all these individuals sign language. There is also an opportunity cost to investing in training for only one impairment type, since this focus would not translate to support for other impairment types (i.e., supporting blind and visually impaired people). Adopting a competency approach—which was not done in most laws—may be a more optimal route for systematically training health workers. For example, in India, the Medical Council of India has included a set of 27 disability competencies in their new curriculum to ensure all doctors are reached with the same skills to serve patients with disabilities. (Singh et al., 2020)

While laws and policies are essential tools for guiding actions and allocating resources, they are most effective when developed in collaboration with affected communities and relevant stakeholders. In many cases, the voices and experiences of disabled people, healthcare professionals, advocacy groups, and other stakeholders are not adequately incorporated into the policymaking process. (Price, 2018) This limited engagement can result in policies that do not fully address the needs and priorities of those directly impacted by disability issues, leading to gaps in implementation and effectiveness. For example, whilst Malawi has made strides in development of policies aimed at addressing the healthcare needs of people with disabilities, challenges such as limited accessibility to healthcare facilities, a high unmet need for assistive technology and insufficient trained personnel remain.(Harrison et al., 2020, Ebuenyi et al., 2023, Munthali et al., 2019) Therefore, future policy development efforts should prioritise inclusive and participatory approaches that ensure the meaningful involvement of all stakeholders throughout the policymaking cycle, from initial design to implementation and evaluation. By fostering greater collaboration and partnership between policymakers, people with disabilities, healthcare providers, and disability advocates, it is possible to develop more comprehensive and responsive policies that better address the complex challenges faced by people with disabilities in accessing healthcare services and achieving health equity.

There is a broader question of whether laws and policies are the appropriate place for such detailed inclusions. Given that these laws and policies were enacted several years ago, it is possible that there are new governments in place, potentially with different priorities that may not include disability-inclusion and disability training for health workers, though many countries in sub-Saharan Africa have had the same political party or president in power for over a decade. (Klobucista, 2023) Laws and policies can be essential to trigger implementation and funding, or may be aspirational, without sufficient actions, meaning that their utility is undermined. For example, Malawi’s documents outlined a clear picture of the training topics, budget, and ministerial responsibilities for accountability. Since this level of commitment and planning was rare in the other reviewed documents, integration of disability competencies into health worker certification may be the more appropriate mechanism for improving health worker competency in these countries. Intervention at this level could enable certified health workers to be trained, examined, and refreshed on their competency in treating disabled patients. Therefore, further research should examine whether these competency, licensing, and Ministry of Health budget documents have greater potential for inclusion of disability than laws. This is particularly important since experimenting with this approach to licensing may trigger more stringent implementation of disability training for health workers. There is a pressing need for further research that delves into the specific provisions and implementations of disability laws and policies related to health worker training globally. Such investigations could offer valuable insights for policymakers, healthcare practitioners, and advocates striving to enhance the inclusivity and effectiveness of healthcare systems across the world.

## Conclusion

Laws, policies, and action plans often signal a country’s intent to change, provide background on specific interventions, or demonstrate values. While mandating, or even including disability training for health workers in laws, policies, and strategies may not necessarily translate into better health outcomes for disabled people, it can be an important step for further action. The policies of many countries in the sub-sample of sub-Saharan African countries have included clauses on disability training, but these lightly recommended actions with limited follow-up plans, strategies, or public funding mean that further work in this area is needed. Policies with specific implementation and monitoring plans need to be developed to financially support and expand health worker training on disability. Examining whether there are licensing requirements related to some form of disability-training would be helpful to understand how countries are implementing these laws.

## Data Availability

Laws and policies are publicly available online.

## Acknowledgements

The authors would like to thank Dr. Emily McFadden, Prof Sara Ryan, and Prof Sue Ziebland for their early input on the framework and preliminary results.

