## Supplementary Table 1 for "Have we forgotten our obligation to train health workers on disability? A policy analysis in sub-Saharan Africa"

| **Country** | **Law/Policy Name (English Translation)** | **Language** | **Date** | **1. Requirements** | **2. Training** | **3. Training Cost** | **4. Training duration** | **5. Competencies covered** | **6. Educational stage** | **7. Training methods** | **8. Impairment-specific** | **9. Cadres** | **10. Benefits for attendance** | **11. Monitoring and Evaluation** |
| --- | --- | --- | --- | --- | --- | --- | --- | --- | --- | --- | --- | --- | --- | --- |
| **Angola** | Lei da Pessoa com Deficiência (Persons with Disabilities Act) Lei n.º 21/12 | Portuguese | 2012 | Recommended |  |  |  |  |  |  |  |  |  |  |
| **Botswana** | National Policy on Care for People with Disabilities | English | 1996 |  |  |  |  |  |  |  |  |  |  |  |
| **Burkina Faso** | Loi no.012 2010/AN du 1er avril 2010, portant protection et promotion des droits des personnes handicapées (Law no.012 2010/AN April 1, 2010, on the protection and promotion of disabled persons) | French | 2010 |  |  |  |  |  |  |  |  |  |  |  |
| **Burkina Faso** | Décret 2012 828 Portant Adoption de Mesures Sociales en Faveur des Personnes Handicapées en Matière de Santé et d'Education (Decree 2012 828 Relative to the Adoption of Social Action in Favor of Disabled Persons in Health and Education) | French | 2012 |  |  |  |  |  |  |  |  |  |  |  |
| **Cabo Verde** | Plano nacional de acção para a década africana das pessoas portadoras de deficiência 2006 2009 (National Plan of Action for the African Decade of Persons with Disabilities 2006 2009) | Portuguese | 2006 | Recommended | Training in specific areas of disability at the level of the main health centres and the training of health professionals are basic conditions for better management of disability issues, making it possible to offer appropriate care and guidance to patients with disabilities. |  |  | Better management of disability issues, enabling appropriate care and guidance to be offered to patients with disabilities. |  |  |  |  |  |  |
| **Cameroon** | Decree No. 2018/6233 fixing the procedures for the application of Law N° 2010/002 of 13 April 2010 on the protection and promotion of persons with disabilities | English | 2018 |  |  |  |  |  |  |  |  |  |  |  |
| **Cameroon** | Loi no.2010/002 du 13 Avril 2010 Portant sur la Protection et la Promotion des Personnes Handicapées (Law no.2010/002 of April 13 2010 on the Protection and Promotion of Disabled Persons) | French | 2010 |  |  |  |  |  |  |  |  |  |  |  |
| **Chad** | [Law No. 007/PR/2007 Bearing Protection for Disabled Persons](https://www.un.org/development/desa/disabilities/wp-content/uploads/sites/15/2019/11/Chad_Law-Bearing-Protection-for-Disabled-Persons-2007.pdf) | French | 2007 |  |  |  |  |  |  |  |  |  |  |  |
| **Congo, Democratic Republic** | Code de la Personne Chapitre III sur les Handicapes, Infirmes et Prodigues (Personal Code Chapter III on the Handicapped, Infirm and Prodigals) | French | 1973 |  |  |  |  |  |  |  |  |  |  |  |
| **Ethiopia** | National Plan of Action of Persons with Disabilities | English | 2012 | Recommended |  |  |  | early identification, service delivery, community based rehabilitation, and family planning and reproductive health services |  |  |  | community based rehabilitation workers; health care personnel in public and private facilities at all levels |  |  |
| **Ethiopia** | National Physical Rehabilitation Strategy | English | 2011 | Recommended |  |  |  | full competency in rehabilitation | CPD |  |  | allied health professionals (physiotherapists, OT) |  |  |
| **Ethiopia** | Initial State Party report to the Committee on the Rights of Persons with Disabilities | English | 2015 |  |  |  |  |  |  |  |  | rehabilitation staff |  |  |
| **Gabon** | Loi no.19/95 Portant organisation de la protection sociale des personnes handicapées au Gabon (Law no.19/95 Relative to the organization of social protection of disabled people in Gabon) | French | 1996 |  |  |  |  |  |  |  |  |  |  |  |
| **Gambia** | Persons with Disabilities Bill, 2020 | English | 2020 | Mandatory |  |  |  |  | Health training institutions' curricula, CPD |  |  | All health and rehabilitation personnel |  |  |
| **Gambia** | Integrated National Disability Policy 2009 2018 | English | 2009 2018 | Recommended |  |  |  |  |  |  |  |  |  |  |
| **Ghana** | [Persons with Disability Act, 2006 (Act 715)](https://www.un.org/development/desa/disabilities/wp-content/uploads/sites/15/2019/11/Ghana_Persons-with-Disability-Act-2006.pdf) | English | 2006 | Mandatory |  |  |  |  | Education programmes |  |  |  |  |  |
| **Kenya** | Initial State Party report to the Committee on the Rights of Persons with Disabilities (2014) | English | 2014 | Mandatory (mainstreamed into health professional curriculum and CPD) |  |  | 3 years (rehab staff) | rehabilitation, early detection, and referrals (CHEWs); some sign language training; information dissemination and rights of people with disabilities | mainstreamed into health professional curriculum and CPD |  |  | rehabilitation professionals (OT, physio, and other service providers), CHEWs, mainstreamed into all health professional curriculum and CPD |  |  |
| **Kenya** | Persons with Disability Act | English | 2003 | Mandatory | acquire skills for proper information dissemination and education on the rights of persons with disabilities. |  |  |  |  |  |  | all healthcare providers |  |  |
| **Lesotho** | National Disability Mainstreaming Plan | English | 2019 | Recommended | Provide sign language skills to health service providers including nurses, doctors, HTC counsellors etc. |  |  | Sign language skills |  |  | Intellectual disabilities and d/Deaf and hard of hearing | Nurses, Doctors, HTC counsellors |  | Plan included |
| **Lesotho** | National Disability and Rehabilitation Policy | English | 2011 | Recommended |  |  |  |  |  |  |  |  |  |  |
| **Madagascar** | Loi sur les personnes handicapees | French | 1998 |  |  |  |  |  |  |  |  |  |  |  |
| **Malawi** | CRPD Initial and Second State Party Reports | English | 2016 |  |  |  |  |  |  |  |  |  |  |  |
| **Malawi** | Disability Act, 2012 | English | 2012 | Mandatory |  |  |  |  |  |  |  |  |  |  |
| **Malawi** | National Disability Mainstreaming Strategy and Implementation Plan 2018 2023 | English | 2018 2023 | Recommended | 3 new curriculums for health personnel; 41 physiotherapists trained; 100 rehabilitation technicians trained; | Free (with 50 million MK budget) |  | Sign language, causes of disability, early intervention, assessment, and referral services | Pre service, CPD |  | Albinism | Physiotherapists, orthopaedics, dermatologists and ophthalmologists, HTC service providers |  | Plan included |
| **Malawi** | National Policy on Equalisation of Opportunities for People with Disabilities, 2006 | English | 2006 | Recommended |  |  |  |  |  |  |  | Trained medial and other rehabilitation personnel |  | Plan included |
| **Mauritania** | Disability Act | English | 2006 |  |  |  |  |  |  |  |  |  |  |  |
| **Mauritania** | Initial State Party report to the Committee on the Rights of Persons with Disabilities | French | 2017 |  |  |  |  |  |  |  |  |  |  |  |
| **Mauritius** | Initial State Party report to the Committee on the Rights of Persons with Disabilities (2014) | English | 2014 |  |  |  |  |  |  |  |  |  |  |  |
| **Mauritius** | National Policy Paper and Action Plan on Disability | English | 2007 | Recommended |  |  |  | disability issues and rehabilitation |  |  |  | Doctors, Paramedics, CBR workers and Social Workers (only existing health workers) |  |  |
| **Mozambique** | Plano Nacional de Acção para Área da Deficiência PNAD II 2012 2019 (National Plan of Action for the Area of Disability PNAD II 2012 2019) | Portuguese | 2012 2019 |  |  |  |  |  |  |  |  |  |  |  |
| **Mozambique** | Estratégia da Pessoa Portadora de Deficiência na Função Pública 2009 2013 (Strategy for persons with disabilities in the public sector 2009 2013) | Portuguese | 2009 2013 |  |  |  |  |  |  |  |  |  |  |  |
| **Namibia** | National Disability Council Act 2004 | English | 2004 | Recommended |  |  |  |  | curriculum |  |  | health workers |  |  |
| **Nigeria** | Discrimination Against Persons with Disabilities Act | English | 2018 |  |  |  |  |  |  |  |  |  |  |  |
| **Nigeria** | Nigerians with Disability Decree | English | 1993 |  |  |  |  |  |  |  |  |  |  |  |
| **Nigeria** | National Policy on Rehabilitation | English | n.d. |  |  |  |  |  |  |  |  |  |  |  |
| **Rwanda** | NCPD Strategic Plan and Its Operational Plan for the Implementation 2013 2018 | English | 2013 2018 |  |  |  |  |  |  |  |  |  |  |  |
| **Rwanda** | Law no.01/2007 Of 20/01/2007 Relating to Protection of Disabled Persons in General | English | 2007 |  |  |  |  |  |  |  |  |  |  |  |
| **Rwanda** | Initial State Party report to the Committee on the Rights of Persons with Disabilities | English | 2015 | Recommended |  |  |  |  | State sector (teachers, police, medical staff, SACCO banks etc.) as a standard part of their qualifying training |  |  |  |  | NCPD Health worker training organized for 600 staff in 30 districts over 3 days Dec 2013 |
| **Senegal** | Initial State Party report to the Committee on the Rights of Persons with Disabilities | French | 2016 |  |  |  |  |  |  |  |  |  |  |  |
| **Seychelles** | Initial State Party report to the Committee on the Rights of Persons with Disabilities | English | 2015 | Recommended |  |  |  |  | National Institute of Health and Social Studies (NIHSS) has required course; child development is done in service |  |  |  |  |  |
| **Sierra Leone** | The Persons With Disability Act, 2011 | English | 2011 |  |  | Contribute to expenses of institutions that train carers of PWDs |  |  |  |  |  |  |  |  |
| **South Africa** | South Africa's National Disability Policy | English | 2015 |  |  |  |  |  |  |  |  |  |  |  |
| **South Africa** | Initial State Party report to the Committee on the Rights of Persons with Disabilities | English | 2015 |  |  |  |  |  |  |  |  |  |  |  |
| **South Africa** | The National Development Plan 2030 Persons with Disabilities as Equal Citizens | English | 2015 | Recommended |  |  |  |  |  | led by resource person with disabilities |  | nurses, doctors, rehabilitation personnel, management and administrative personnel |  |  |
| **South Africa** | National Rehabilitation Policy | English | 2000 | Recommended |  |  |  |  |  |  |  |  |  |  |
| **Tanzania** | The Persons with Disability Act | English | 2011 | Mandatory | increase knowledge, disability sensitive awareness, respect for rights, dignity, and needs of persons with disabilities. |  |  |  |  |  |  | all health and rehabilitation personnel |  |  |
| **Tanzania** | National Policy on Disability 2008 | English | 2008 | Recommended |  |  |  |  |  |  |  | "all personnel in service delivery" |  |  |
| **Togo** | Loi Togolaise Relative a La Protection Sociale des Personnes Handicapees | French | 2011 | Recommended |  |  |  |  |  |  |  |  |  |  |
| **Uganda** | Persons with Disabilities Act | English | 2006 | Mandatory | sign language |  |  | sign language |  |  |  |  |  |  |
| **Uganda** | National Council for Disability Act | English | 2003 |  |  |  |  |  |  |  |  |  |  |  |
| **Uganda** | Initial State Party report to the Committee on the Rights of Persons with Disabilities | English | 2015 | Mandatory (mainstreamed into health professional curriculum) | p 39 The Government has implemented several programmes including distribution of assistive devices like wheelchairs and prevention of blindness. Aspects of disability and managing disability from the social perspective are being included into the training curriculum of health workers. p 40: establish medical rehabilitation departments or sections in hospitals, special institutions of rehabilitation and carry out clinical practice and training, scientific research, personnel training and work of technical guidance in the field of rehabilitation; provide various forms of technical training... for personnel engaged in rehabilitation work |  |  |  |  |  |  |  |  |  |
| **Zambia** | The Person with Disability Act 2012 | English | 2012 | Mandatory |  |  |  |  | in curricula |  |  |  |  |  |
| **Zimbabwe** | National Disability Policy (https://www.veritaszim.net/sites/veritas_d/files/National%20Disability%20Policy%20V4%28White%20Background%29.pdf) | English | 2021 | Mandatory |  |  |  |  |  |  |  |  |  |  |
